# Pesticide Issue Interest and Related Factors Among Japanese University Students: An Environmental Health Risk Perception Perspective Focusing on Neonicotinoids

**DOI:** 10.64898/2026.01.07.25343027

**Authors:** Yuusuke Harada, Keiko Nosaka, Michiko Miyakawa

## Abstract

**Background:** Neonicotinoid pesticides are systemic insecticides with neurotoxic potential and environmental persistence, raising concerns about chronic low-dose exposure in humans and impacts on ecosystems. In Japan, regulatory restrictions are generally less stringent than in the European Union, yet risk perception among young adults remains underexplored.

**Objective:** To examine factors associated with university students’ interest in pesticide-related issues, focusing on family discussions about environmental topics, household preference for organic vegetables, agricultural experience, and depressive symptoms.

**Methods:** We conducted a cross-sectional questionnaire survey among 690 undergraduate students at a private university in the Tokyo metropolitan area (academic year 2024). Interest in pesticide issues was assessed on a 4-point scale and dichotomized as high (very/somewhat) vs low (not very/not at all). Explanatory variables included frequency of family discussions about environmental issues, household preference for organic vegetables, agricultural experience, the Quick Inventory of Depressive Symptomatology–Self Report (QIDS-SR-J), age, and sex. Multivariable logistic regression was used to estimate adjusted odds ratios (aORs) and 95% confidence intervals (CIs).

**Results:** Participants’ mean age was 20.6 (SD 4.9) years and mean QIDS-SR-J score was 6.0 (SD 4.2). Overall, 74.6% reported high interest in pesticide issues. High interest was associated with more frequent family discussions about environmental issues (aOR 2.22, 95% CI 1.50–3.30; p<0.001) and household preference for organic vegetables (aOR 1.73, 95% CI 1.14–2.62; p=0.009). Agricultural experience, depressive symptoms, age, and sex were not significantly associated. Notably, while 53.2% supported pesticide-free/organic farming, 85.5% avoided blemished (insect-damaged) vegetables and 77.1% considered pesticides necessary, suggesting an attitude–behavior gap.

**Conclusions:** Among Japanese university students, interest in pesticide issues was linked to family-level environmental discourse and household organic orientation, rather than depressive symptoms. The observed gap between pro-organic attitudes and consumer preferences underscores the need for risk communication and environmental health education that connect values to feasible purchasing behaviors.

## Introduction

Neonicotinoid pesticides have been widely used since the 1990s as systemic insecticides targeting nicotinic acetylcholine receptors. Although they provide strong insecticidal efficacy, concerns have been raised about potential effects on non-target organisms, including mammals, as well as their environmental persistence and mobility [1,2].

In humans, accumulating evidence suggests potential associations between low-dose chronic exposure and neurodevelopmental outcomes, cognitive or behavioral changes, and cardiovascular effects [2,3]. Multiple exposure routes have been reported, including dietary intake through agricultural products, household insecticides, pet products, termite control treatments, indoor air, and house dust [4,5]. Biomonitoring studies in Japan have detected several neonicotinoids and metabolites in urine samples from adults and children, indicating ubiquitous background exposure [2,6].

Ecological concerns include impacts on pollinators, aquatic invertebrates, and birds, with implications for ecosystem services such as pollination and pest control. In response to these concerns, the European Union implemented restrictions including outdoor use bans for several neonicotinoids, informed by the precautionary principle [7,8]. In contrast, pesticide regulation and residue standards in Japan are often considered less restrictive, prompting debate on how to communicate risks and benefits and how to design effective policies [4,5,9,10].

Risk perception regarding food safety and environmental hazards is shaped by information environments, education, family conversations, and personal values [11–13]. Prior work in Japan has noted a gap between positive attitudes toward organic products and actual purchasing behavior, influenced by factors such as price, appearance, and availability [14–17].

University students represent both current consumers and future professionals and voters who will participate in decisions about environmental health risks. This study therefore examined correlates of interest in pesticide-related issues among Japanese university students, with particular attention to family discussions on environmental issues, household organic preferences, agricultural experience, and depressive symptoms.

### Methods

### Study design and participants

This was a cross-sectional questionnaire survey conducted among undergraduate students enrolled in a course at a private university in the Tokyo metropolitan area during the 2024 academic year. Paper-based anonymous questionnaires were distributed and collected during class time. A total of 690 students provided valid responses and were included in the analysis.

### Ethics approval and consent

The study protocol was approved by the Ethics Committee of Hosei University (Approval No.: Graduate School/Public Health Ethics 2403). Participants were informed verbally and in writing about the study purpose, voluntary participation, anonymity, and publication of aggregated results. Only students who provided consent were included.

### Measures

Sociodemographic variables included age and sex.

Depressive symptoms were assessed using the Japanese version of the Quick Inventory of Depressive Symptomatology–Self Report (QIDS-SR-J; score range 0–27) [18–20]. The total score was used as a continuous variable; severity categories were defined as 0–5 (none), 6–10 (mild), 11–15 (moderate), 16–20 (severe), and 21–27 (very severe).

Interest in pesticide issues was assessed by the question, “How interested are you in pesticide-related issues?” with four response options (very interested, somewhat interested, not very interested, not at all interested). Responses were dichotomized as high interest (very/somewhat) vs low interest (not very/not at all).

Family discussion about environmental issues was assessed on a four-point scale (often, sometimes, rarely, never) and dichotomized as higher frequency (often/sometimes) vs lower frequency (rarely/never).

Household preference for organic vegetables was assessed (yes/no) as whether the household tried to choose pesticide-free or organic vegetables.

Agricultural experience was assessed (yes/no) as having participated in farming experiences through school or community activities.

Attitudes toward pesticide-free/organic farming, avoidance of blemished (insect-damaged) vegetables, and perceived necessity of pesticides were assessed using three items, as described in Tables 2–3.

**Table 1.** Participant characteristics and related factors (N = 690)

| Measure/Item | Category | n | % or value |
| --- | --- | --- | --- |
| Age (years) | — | — | 20.6 ± 4.9 |
| QIDS-SR-J score | — | — | 6.0 ± 4.2 (range 0–21) |
| QIDS-SR-J severity | None (0–5) | 368 | 53.3 |
|  | Mild (6–10) | 231 | 33.5 |
|  | Moderate (11–15) | 67 | 9.7 |
|  | Severe (16–20) | 23 | 3.3 |
|  | Very severe (21–27) | 1 | 0.1 |
| Sex | Male | 309 | 44.8 |
|  | Female | 378 | 54.8 |
|  | Prefer not to say | 3 | 0.4 |
| Family discussion about environmental issues | Often | 35 | 5.1 |
|  | Sometimes | 241 | 34.9 |
|  | Rarely | 296 | 42.9 |
|  | Never | 118 | 17.1 |
| Household organic preference | Yes (tries to choose pesticide-free/organic vegetables) | 217 | 31.4 |
|  | No | 473 | 68.6 |
| Agricultural experience | Yes | 578 | 83.8 |
|  | No | 112 | 16.2 |
| Interest in pesticide issues | Very interested | 82 | 11.9 |
|  | Somewhat interested | 433 | 62.8 |
|  | Not very interested | 164 | 23.8 |
|  | Not at all interested | 11 | 1.6 |

**Table 2.** Support for organic farming and avoidance of blemished (insect-damaged) vegetables.

| Support for pesticide-free/organic farming | Does not avoid blemished vegetables | Avoids blemished vegetables | Row total |
| --- | --- | --- | --- |
| Do not support / undecided | 44 (13.6%) | 279 (86.4%) | 323 |
| Support | 56 (15.3%) | 311 (84.7%) | 367 |

### Statistical analysis

Descriptive statistics were calculated as mean (standard deviation) or n (%). We cross-tabulated support for pesticide-free/organic farming with (i) avoidance of blemished vegetables (Table 2) and (ii) perceived necessity of pesticides (Table 3).

**Table 3.**
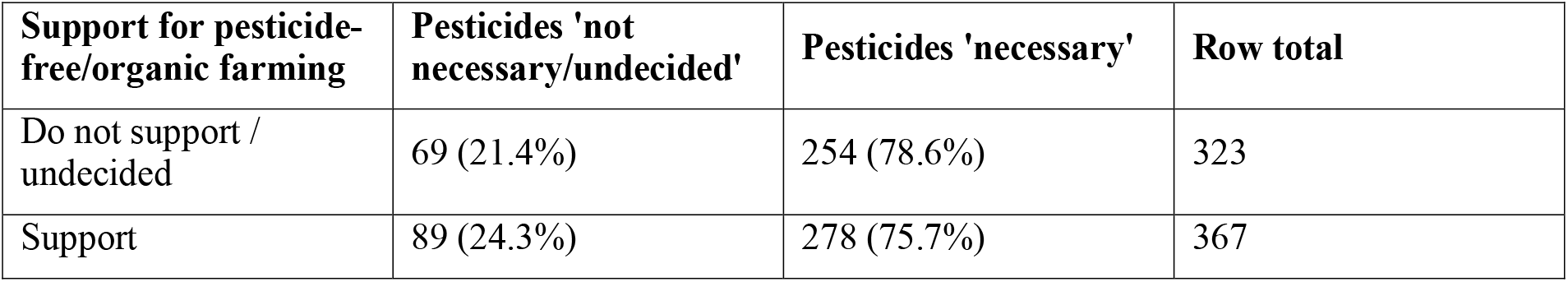
Support for organic farming and perceived necessity of pesticides.

To identify correlates of high interest in pesticide issues, we fitted a multivariable logistic regression model with high interest as the dependent variable (1=high, 0=low). Explanatory variables were frequency of family environmental discussions, household preference for organic vegetables, agricultural experience, QIDS-SR-J score, age, and sex (female=1, male=0). Adjusted odds ratios (aORs) and 95% CIs were reported. Statistical significance was set at two-sided p<0.05. Analyses were performed using Python (version 3.13).

## Results

### Participant characteristics

Table 1 summarizes participant characteristics and key variables. The mean age was 20.6 (SD 4.9) years. The mean QIDS-SR-J score was 6.0 (SD 4.2), with 53.3% classified as none and 33.5% as mild depressive symptoms. Overall, 40.0% reported that environmental issues were often or sometimes discussed in their families, and 31.4% reported a household preference for organic vegetables. Agricultural experience was reported by 83.8% of participants. High interest in pesticide issues was reported by 74.6% of participants.

### Support for organic farming and consumer preferences

Over half of participants (53.2%) supported pesticide-free/organic farming. However, 85.5% reported avoiding blemished (insect-damaged) vegetables. As shown in Table 2, avoidance of blemished vegetables was common regardless of support for organic farming.

Regarding perceived necessity of pesticides, 77.1% responded that pesticides were necessary or somewhat necessary. Table 3 shows that approximately three-quarters of participants in both groups (support vs no/undecided) perceived pesticides as necessary, suggesting a gap between ideological support and practical preferences.

### Correlates of high interest in pesticide issues

In multivariable logistic regression (Table 4), high interest in pesticide issues was significantly associated with higher frequency of family discussions about environmental issues (aOR 2.22, 95% CI 1.50–3.30; p<0.001) and household preference for organic vegetables (aOR 1.73, 95% CI 1.14–2.62; p=0.009). Agricultural experience, QIDS-SR-J score, age, and sex were not statistically significant predictors.

**Table 4.** Multivariable logistic regression for high interest in pesticide issues (N = 690)

| Variable | Category / unit | aOR | 95% CI | p-value |
| --- | --- | --- | --- | --- |
| Family discussion about environmental issues | Higher frequency (often/sometimes) vs lower (rarely/never) | 2.22 | 1.50–3.30 | <0.001 |
| Household organic preference | Yes vs no | 1.73 | 1.14–2.62 | 0.009 |
| Agricultural experience | Yes vs no | 1.28 | 0.80–2.04 | 0.299 |
| QIDS-SR-J score | Per 1-point increase | 0.99 | 0.95–1.03 | 0.608 |
| Age | Per 1-year increase | 1.03 | 0.98–1.09 | 0.190 |
| Sex | Female vs male | 1.42 | 0.99–2.02 | 0.055 |

Abbreviations: aOR, adjusted odds ratio; CI, confidence interval; QIDS-SR-J, Quick Inventory of Depressive Symptomatology–Self Report, Japanese version.

## Discussion

This cross-sectional study found that Japanese university students’ interest in pesticide-related issues was associated with family-level environmental discourse and household organic orientation, but not with depressive symptoms or agricultural experience. Depressive symptoms were included as an exploratory covariate because affective states can influence risk perception [11], and epidemiological syntheses have reported associations between pesticide exposure or poisoning and depressive symptoms in agricultural populations [22,23]. These results highlight the role of household socialization and values in shaping environmental health risk perception among young adults.

The attitude–behavior gap observed in this study was notable: while more than half supported pesticide-free/organic farming, most participants avoided blemished vegetables and considered pesticides necessary. This pattern aligns with prior research indicating that purchasing decisions are driven by multiple competing factors such as appearance, convenience, and price, and that pro-environmental attitudes do not automatically translate into behavior [11,16,17].

From a public health and environmental policy perspective, these findings suggest that education and risk communication should go beyond emphasizing hazards. Practical communication strategies may need to address perceived barriers, provide balanced information about risks and benefits, and connect consumers with feasible options (e.g., labeling, access, and affordability initiatives).

Although neonicotinoids have been linked to potential human and ecological risks, regulatory approaches differ across regions. The precautionary principle has shaped EU restrictions [7,8], whereas Japan’s regulatory environment has been characterized as comparatively permissive [4,5]. Understanding young adults’ risk perception can inform future policy debates and educational interventions.

### Limitations

This study has limitations. First, participants were recruited from a single university, which limits generalizability. Second, all measures relied on self-report and may be subject to social desirability or recall bias. In particular, agricultural experience was assessed with a single yes/no item and may have occurred years earlier; we did not capture timing, frequency, or intensity, so misclassification due to imperfect recall is possible. Third, we did not measure pesticide exposure biomarkers or dietary intake; therefore, relationships between risk perception and actual exposure remain unknown. Fourth, depressive symptom severity was generally low in this sample, limiting power to detect associations with risk perception.

## Conclusions

Interest in pesticide-related issues among Japanese university students was associated with family discussions about environmental issues and household preference for organic vegetables, but not with depressive symptoms. The observed attitude–behavior gap indicates a need for environmental health education and risk communication that bridge values and feasible consumer choices, alongside broader policy discussions grounded in the precautionary principle.

## Declarations

### Ethics approval and consent to participate

Approved by the Ethics Committee of Hosei University (Approval No.: Graduate School/Public Health Ethics 2403). Written informed consent was obtained from all participants.

### Consent for publication

Not applicable (no identifiable individual data are presented).

### Data availability

The dataset contains questionnaire responses collected under ethical approval. De-identified data may be made available by the corresponding author upon reasonable request and subject to institutional approval.

### Authors’ contributions

Yuusuke Harada: conceptualization, formal analysis, writing - original draft. Keiko Nosaka: data collection, writing - review & editing. Michiko Miyakawa: supervision, writing - review & editing.

### Reporting guideline

This manuscript follows the STROBE (Strengthening the Reporting of Observational Studies in Epidemiology) checklist for cross-sectional studies; the completed checklist is provided as a supplementary file.

## Acknowledgements

We thank the participating students and course staff for their cooperation.

